# Inherent random fluctuations in COVID-19 outbreaks may explain rapid growth of new mutated virus variants

**DOI:** 10.1101/2021.01.07.21249353

**Authors:** Kenneth Bodin, Joacim Rocklöv

## Abstract

A new virus variant of SARS-COV-2 has had a profound impact on society while governments have taken action to limit its impacts by enforcing lockdowns and limiting spread from the UK to other countries. Variants with mutations in the virus genome are likely to occur, but do not always associate to significant changes in the biology of the virus, or the disease. For the variant VOC 202012/01 (also referred to as B.1.1.7), however, preliminary reports indicate it may be more transmissible. Here we use a simulation model calibrated to the inherent random fluctuating transmission pattern of COVID-19 to investigate what the probability may be for detecting more transmissible virus variants post facto. We find that post facto identification of successful virus variants of SARS-COV-2 are likely to exhibit growth rates that are substantially larger than the average growth rate. This finding has implications for interpreting growth rate and transmissibility of new virus variants.

## Main text

On December 14, authorities in the United Kingdom reported that a new mutated variant of SARS-CoV-2 had been sequenced, referred to as VOC 202012/01 or B.1.1.7. The new variant could make preventive actions to suppress the COVID-19 pandemic much harder (until effective vaccines have been rolled out), if the preliminary reports of increased transmissibility are correct (1). Therefore, many governments have implemented policies to avoid importing, and to slow the importations of the new virus variant to their country, such as canceling flight traffic, testing and quarantine of air travellers from the UK, border closure for freight transports and trucks, and increased local efforts to survey the circulation and prevalence of virus strains.

Behind triggering the alarm stands both biological and epidemiological insights and data. The SARS-COV-2 variant VOC 202012/01 has 17 mutations, some of potential suspected biological significance linked to its transmissibility (2). Complementary data analysis indicates that this variant could be 56% (2) or 71% (3) more transmissible, and likewise proportionally affect the reproduction number, R. Other reports indicate an increase in R in the range of 0.39-0.93 (3).

Without excluding any biological changes of significance affecting transmissibility, we provide here a potential alternative explanation of the observed patterns of the new virus variants, such as VOC 202012/01, when they are observed dominant in retrospect. Due to the seemingly random inherent fluctuating patterns of the transmission of SARS-CoV-2, we can illustrate using simulations that post-facto discovered rapid growth of new variants may well be explained by the heterogeneous reproduction that characterizes SARS-CoV-2. In fact, we show that specific variants of the virus that survive for many generations are likely to be associated with higher Re and more dominance due to a majority of variants ending by stochastic extinction. We find that only a smaller proportion of biologically equal variants manage to survive up to 10 generations, while most of them die out early. The difference between survival and extinction can be an early advantageous stochastic event, such as a superspreading.

A recent Chinese study with rigourous contact tracing indicates that 15% of all primary infections of SARS-CoV-2 caused 80% of all secondary infections (5). This has also previously been seen in contact tracing data (6), in genome viral sequencing data (7), and in epidemiological data analysis (8). The individual reproductive numbers (i.e. the number of secondary cases originating from one infection) associated with such inhomogeneous spreading patterns can be described well by a heavy tail distribution, typically the negative binomial distribution (5,9). While the individual reproduction number measures the individual inhomogeneous patterns of transmission, the R value measures the average number over all individual secondary infections in a population.

Inspired by Althouse et.al. (10) we apply a branching model with a negative binomial distribution (NB). We set the effective reproductive number of the NB, R=1.3, to represent the current range of the recent R in the UK (1). We use a dispersion parameter of k=0.30, taken from Sun et al (5) representing a conservative suppression situation. The generation time is not of large importance to our simulation, but has been estimated to around 5.3 days. However, it can be shorter with effective case isolation and longer with ineffective case isolation (5). From this parameterisation, we grow the transmission chains trajectories iteratively. The index case is one infected individual, which infects a new number of individuals according to a random number generated from the NB distribution. This yields the number of infected in the second generation. For each individual infected in this second generation, we draw a new random number for the third generation, and so on. Henceforth, we establish the full transmission chain from each index case up to generation 10 and we refer to these as individual transmission chain trajectories. Assuming only one virus variant is circulating, these individuals will all be infected by the same virus variant with the same biological characteristics and transmissibility. Repeating this process a large number of times can help us understand what the natural variability of the transmission trajectories are. In fact, the experiment will allow us to estimate how often the Re value of a trajectory, i.e., virus variant, would be higher than the average Re value of known virus variants by pure chance. It thus serves as an important natural control to any data driven investigation of detecting important new alternations of Re, or i.e., transmissibility, of new virus genetic variants.

We simulate 10 generations, corresponding to approximately 53 days, and estimate transmission chain trajectories for 100.000 infected index individuals carrying the same virus variant. We run the branching processes simulation and track each transmission chain following from each index case for the 10 generations. We assume that immunity is already accounted for in R and does not further affect the reproduction during the simulation. Figure 1 illustrates all transmission chain trajectories and the number of individuals being infected from each index case up to generation 10. We observe that many of the transmission chains exhibit stochastic extinction. The average Re of the surviving trajectories at generation 10 is 1.6. This is approximately 23% higher than the actual R of the NB. Using a constant, deterministic number of infections generated in each generation of Re equal to 1.7, it closes in and equals the average of the surviving trajectories of R=1.3, first at generation 8.

**Figure 1.**
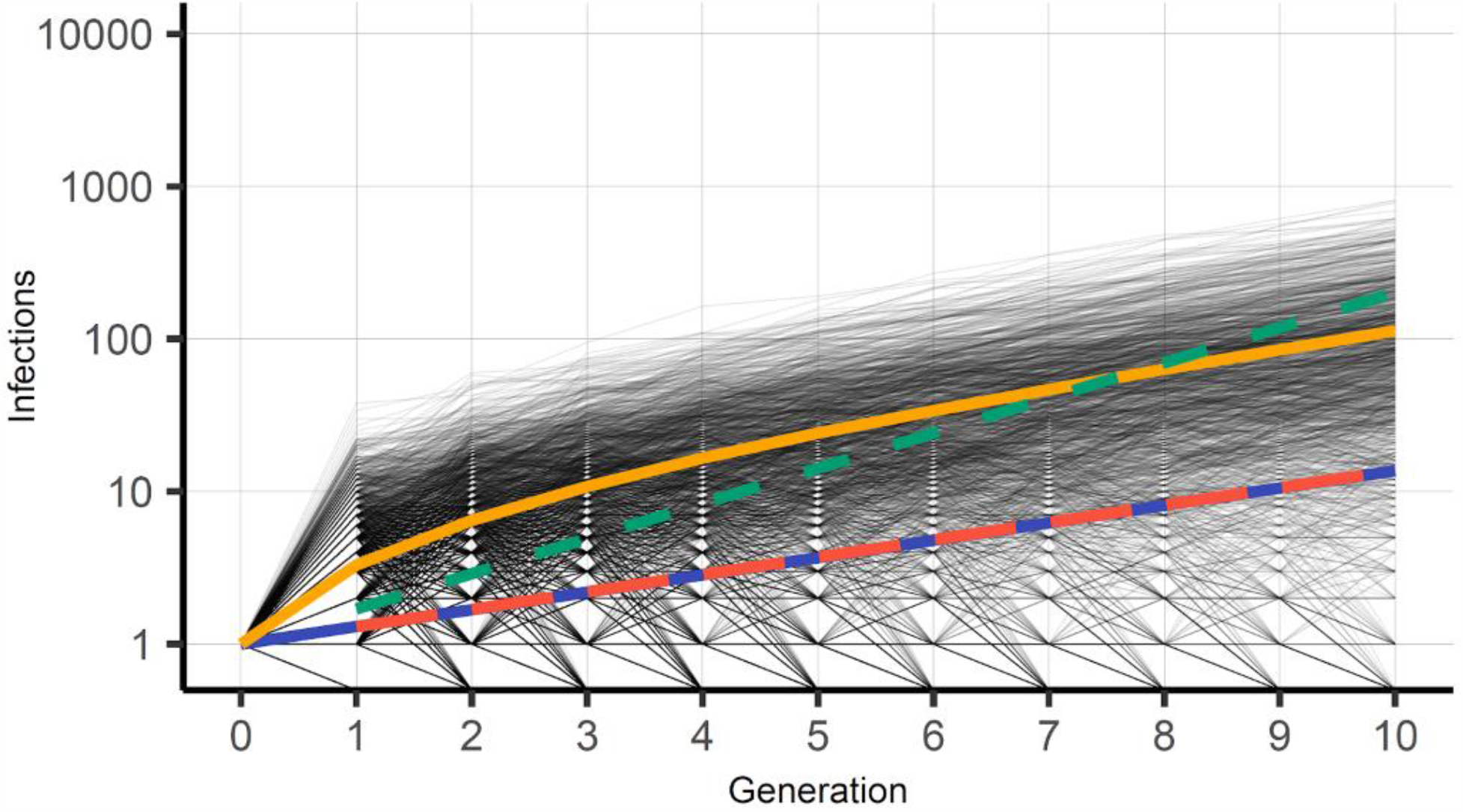
The trajectories of new infections at each generation resulting from the 100,000 index infections and biologically similar strains at generation 0 (grey lines). The yellow line shows the average number of infections among the trajectories that are sustained (only 12.1% in generation 10). The blue line shows the deterministic number of infections from R equal to 1.3 and the dashed red line shows the corresponding average measured in the simulation. The green dashed line shows the deterministic number of infections if R is equal to 1.7, which crosses the average of surviving trajectories of Re 1.3 first at generation 8.

In Figure 2, we show that a high number of strains from the 100,000 index infections die out before the 10th generation, in fact, as many as 87.9% go extinct before or at generation 10. This is well known to occur in real life situations, but the frequency of occurrence is related to the stochastic superspreading pattern of SARS-COV-2 and sensitive to the parameter k and the R (11). We find that as many as 20.4% of the surviving strains could be expected to have an Re of 1.7 or above, substantially higher than the average Re of 1.3 for all trajectories. On the other hand, a virus variant with even higher transmission, say an Re of 2, would generate 1024 infections in generation 10 of Figure 1, and would be an outlier in the histogram of Figure 2 with a probability as small as 0.024%. We note, however, that the difference to the average surviving trajectories average 1.6 is much less than to the overall average of the NB distribution of 1.3.

**Figure 2.**
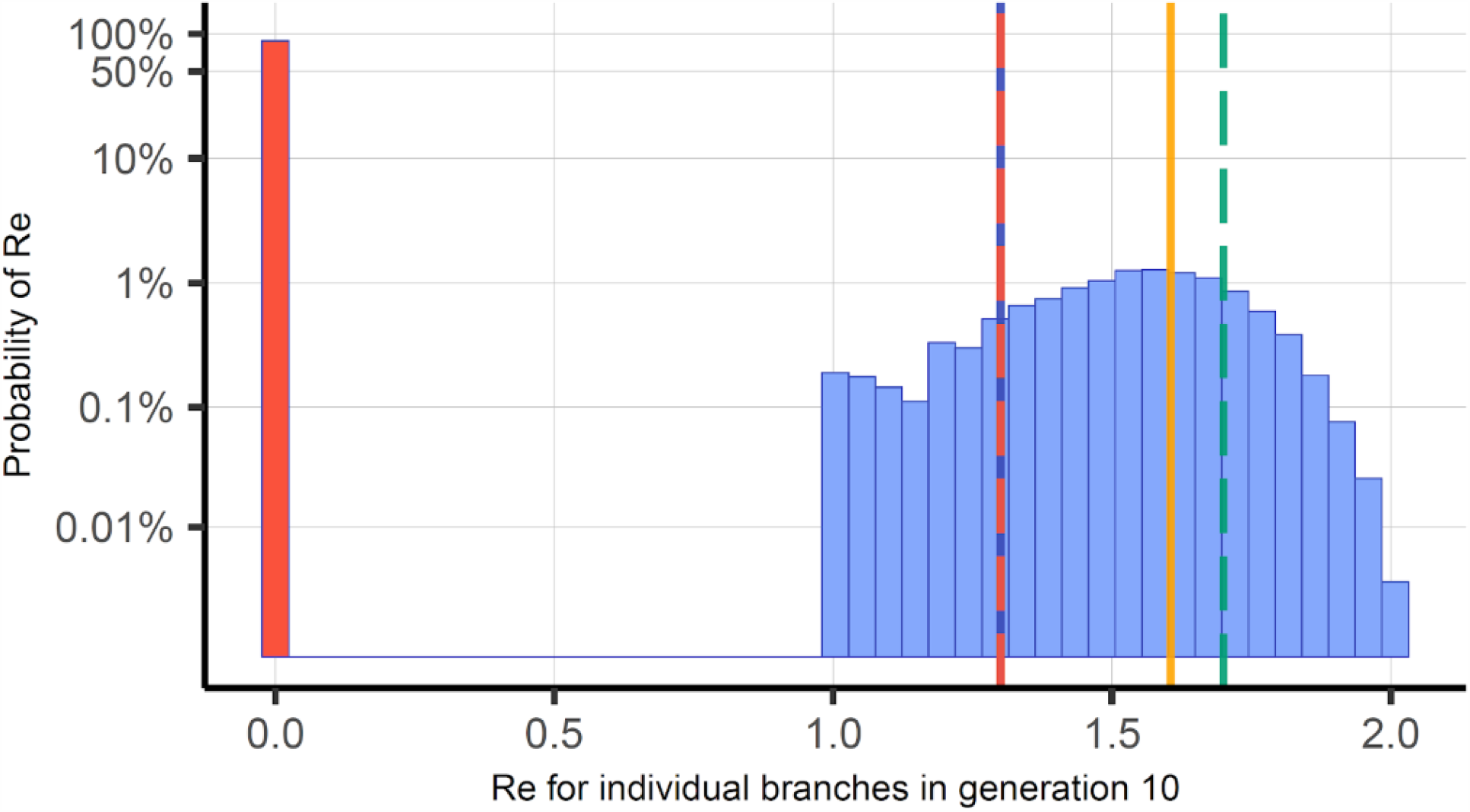
The frequency of the Re’s of the different transmission chain trajectories after 10 generations. The strains that go extinct are described by the red bar and equals 87.9%. The remaining Re estimates fall between 1.0 to 2.0. Re of 1.3 is marked by the red and blue vertical line, the average Re of the surviving transmission chains is 1.6 and marked by the vertical yellow line. 20.4% of the surviving transmission chains have Re values above 1.7 by random (dashed green line).

When we change the parameter k to 0.15, corresponding to a dispersion parameter of a non-suppressed outbreak (7), the extinction rate in generation 10 increases to 92.1% and the probability of a surviving trajectory with Re greater than 1.7 becomes 34.9%. The probability for a trajectory with Re greater than 2.0 increases to 1.4%. Thus, the effect from random fluctuations is expected to be even more prominent in communities with a lower degree of non-pharmaceutical interventions or lower compliance to regulations, in contrast to China where COVID-19 was effectively suppressed.

Our simulation experiment indicates, given that the modelling assumptions would hold in reality, that there is a probability that any post-facto observed virus variant, such as the VOC 202012/01, can in fact be the product of just a very successful random transmission chain trajectory without alternations in the transmissibility, at least this is probable to happen by chance to some degree. Here we show that 20.4% of the transmission trajectories show at least an Re shift of +0.4 higher than the average. It is important to adjust for this in future studies of changes in transmissibility and Re values of new virus variants, such as VOC 202012/01. This said, the simulations indicate that the longer the generations are run, pattern stagnates, and this may be less of a problem in situations where the number of generations observed are very large. Finally, we want to stress that we are not providing any empirical proof in any way that the VOC 202012/01 is not more transmissible, it may well be, but we do show that there is a bias when observing post facto successful virus variants, which need to be adjusted for in any data analysis of transmissibility. This has bearing for emerging strains and policy implications for COVID-19, as well as for other infectious diseases, especially those exhibiting substantial inhomogeneous spreading patterns.

## Data Availability

Data from computer simulations.

